# A signature of pre-operative biomarkers of cellular senescence to predict risk of cardiac and kidney adverse events after cardiac surgery

**DOI:** 10.1101/2023.04.03.23288081

**Authors:** Amy Entwistle, Susan Walker, Anne Knecht, Susan L. Strum, Asad Shah, Mihai V. Podgoreanu, Aliaksei Pustavoitau, Natalia Mitin, Judson B. Williams

**Author notes:** Co-corresponding authors: Judson Williams, MD and Natalia Mitin, PhD.

## Abstract

**Objective:** Understand the potential for pre-operative biomarkers of cellular senescence, a primary aging mechanism, to predict risk of cardiac surgery-associated adverse events.

**Methods:** Biomarkers of senescence were assessed in blood samples collected prior to surgery in 331 patients undergoing CABG +/-valve repair or replacement. Patients were followed throughout the hospital stay and at a 30-day follow-up visit. Logistic regression models for pre-operative risk prediction were built for age-related clinical outcomes with high incidence including KDIGO-defined acute kidney injury (AKI), decline in eGFR ≥25% between pre-op and 30 days, and MACKE30, a composite endpoint of major adverse cardiac and kidney events at 30d.

**Results:** AKI occurred in 19.9% of patients, persistent decline in kidney function at 30d occurred in 11.0%, and MACKE30 occurred in 13.4%. A network of six biomarkers of senescence (p16, p14, LAG3, CD244, CD28 and suPAR) were able to identify patients at risk for AKI (AUC 0.76), kidney decline at 30d (AUC 0.73), and MACKE30 (AUC 0.71). Comparing the top and bottom tertiles of senescence-based risk models, patients in the top tertile had 7.8 (3.3-8.4) higher odds of developing AKI, 4.5 (1.6-12.6) higher odds of developing renal decline at 30d, and 5.7 (2.1-15.6) higher odds of developing MACKE30. All models remained significant when adjusted for clinical variables. Patients with kidney function decline at 30d were largely non-overlapping and clinically distinct from those who experienced AKI, suggesting a different etiology. Typical clinical factors that predispose to AKI (e.g., age, CKD, surgery type) associated with AKI but not the 30d decline endpoint which was instead associated with new-onset atrial fibrillation.

**Conclusions:** A six-member network of biomarkers of senescence, a fundamental mechanism of aging, can identify patients for risk of adverse kidney and cardiac events when measured pre-operatively.

## INTRODUCTION

Aging confers risk for most medical conditions. Cellular senescence is a well-known aging mechanism that links deleterious subcellular changes with multi-system loss of organ function and physiologic decline. Senescent cells are permanently growth arrested but metabolically active, secreting pro-inflammatory and pro-fibrotic cytokines that contribute to chronic inflammation and impaired tissue regeneration ^1, 2^.

These kinds of aging-related vulnerabilities can manifest as adverse events after major medical interventions. Such risks may be discordant with expectations based on chronological age and multi-morbidities. The latter are routinely considered in medical decision making and form the foundation of pre-surgical risk assessment. In the surgical setting, measuring aging-related risk pre-operatively through molecular biomarkers could allow for optimization of care pathways across the peri-operative period.

Cardiac surgery has been at the forefront of risk mitigation and outcomes reporting, including quality improvement initiatives such as ERAS Cardiac^3^. However, post-operative morbidity and mortality after coronary artery bypass grafting (CABG), the most common form of cardiac surgery, remain high. Older, more co-morbid patients, increasing procedural costs, and value-based payments are driving strong interest in accurate, pre-operative patient stratification and targeted risk mitigation for cardiac patients. A better understanding of aging-related patient risk remains an unexplored area, ripe for discovery and integration into clinical practice. In this study we report the use of network of cellular senescence biomarkers to identify patients at-risk of cardiac surgery-associated adverse events.

## METHODS

### Study design and participants

In the pilot study, adult patients >50 years and older undergoing primary elective or urgent, on-pump, CABG (isolated or combined with valve surgery) were prospectively enrolled at the Johns Hopkins University Hospital between September 2010 and March 2013, or Duke University Hospital between June 2015 and July 2017.

Patients were excluded if they required emergency or salvage CABG, aortic aneurysm or congenital heart disease repair, primary ventricular assist device implantation, severe heart failure (LVEF <25%), hemodynamic instability requiring preoperative vasopressors or IABP, pre-existing end stage kidney disease (eGFR <15 mL/min/1.73m^2^) or renal transplantation, chronic liver disease or cirrhosis, or presence of major active infection (chronic or acute e.g., sepsis, HIV, pneumonia). A total of 46 and 42 participants were enrolled from Duke University and Johns Hopkins Hospitals, respectively.

In the GUARD-AKI study, adult patients (>40 years) undergoing non-emergency (urgent or scheduled) cardiac surgery using cardiopulmonary bypass (CABG or combined CABG/valve) were prospectively enrolled at WakeMed Health and Hospitals, Johns Hopkins University Hospital, and Hoag Memorial Hospital Presbyterian between October 2020 and July 2022. Patients were excluded if they required emergency or salvage CABG, off-pump coronary bypass grafting, aortic aneurysm or congenital heart disease repair, primary ventricular assist device implantation, severe heart failure (LVEF <25%), hemodynamic instability requiring preoperative vasopressors or IABP, pre-existing end stage kidney disease (eGFR <30 mL/min/1.73m^2^) or renal transplantation, chronic liver disease or cirrhosis, or presence of major active infection (chronic or acute e.g., sepsis, HIV, pneumonia). All participants were tested for COVID infection per standard of care in preparation for surgery. This clinical study was registered with clinicaltrials.gov (NCT03635606). IRB of Johns Hopkins University, Duke University, and Central IRB (WIRB/WCG) overseeing WakeMed Health and Hospitals and Hoag Memorial Hospital Presbyterian gave ethical approval for this work.

Power calculations for the GUARD-AKI study were based on the pilot study for a primary endpoint of AKI. The number is predetermined to achieve a 95% confidence interval width of 0.175% on the area under the curve (AUC) of the receiver operating curve (ROC) given that events to non-events occur in a 1:4 ratio such that: a random sample of 40 subjects from the AKI positive population and 158 subjects from the AKI negative population produce a two-sided 95.0% confidence interval with a width of 0.175 when the sample AUC is 0.86. As such the lower limit of the 95% CI is 0.776 and the upper limit is 0.947. Upon study conduct, it was noted that the event to non-event ratio was 1:5 and hence enrollment was increased to achieve the minimum number of required events.

#### Outcome definitions

Patients were followed for the duration of their hospital stay and at 30-day follow-up with the surgeon. In all cohorts, the primary endpoint was development of stage 1 or higher postoperative AKI as defined by KDIGO (sCr increase of ≥0.3 mg/dL in the first 48h or a relative increase of ≥50% in peak sCr from baseline within 7 days post-surgery). GUARD-AKI had additional outcome measures: development of worsening renal function (a ≥25% reduction from baseline eGFR at the 30-day follow-up visit); development of 30-day major adverse kidney events (MAKE30): a composite of persistently impaired renal function (a ≥25% reduction from baseline eGFR at the 30-day follow-up visit, new dialysis, and death); development of 30-day major adverse cardiac events (MACE30): a composite of myocardial infarction (MI), stroke, heart failure, and death; and development of the combination of MAKE30 and MACE30 (major adverse cardiac and kidney events [MACKE30]). A creatinine-based eGFR equation that does not incorporate race, developed by CKD-EPI ^4^, was used to calculate eGFR for all participants.

#### Sample collection and biomarker measurements

Peripheral blood samples were collected prior to surgery, stabilized according to Sapere Bio protocol and processed either on site (Duke University or Johns Hopkins University) or at Sapere Bio. 7.5ml of stabilized blood was used to isolate T cells as previously reported ^5^ and the remainder of the sample was used to isolate plasma. Samples were stored at -80°C until the analysis. Gene expression of p14, p16, LAG3, CD28, and CD244 was analyzed by real-time qPCR. Expression for each gene was normalized to expression of a housekeeping gene. Positive and negative controls were included in each run, and Cts over 37 were considered below the limit of detection. Expression of each senescence gene is reported in log2 units and as arbitrary units, as is standard for qPCR reporting of gene expression levels.

Expression of suPAR, sTNF-R1, and Activin A was measured by ELISA in plasma using commercial kits (R&D Systems) per manufacturer’s instructions. In all analyses, lab personnel were blinded to clinical information and outcome measures.

#### Statistical analyses

*Pilot Study—*To build a predictive model for AKI, expression of p16, p14 (and their second-degree interaction), pre-operative sCr, as well as demographic and clinical variables such as gender, diabetes, and surgery type were tested in a logistic model. Factors for the logistic model for each outcome were chosen by backwards elimination. Akaike information criterion with correction for sample size (AICc) was used to find the model with both good fit to the truth and few parameters.

*GUARD-AKI –* Descriptive statistics were reported as mean (sd) for continuous variables, and as frequency (percentage) for categorical variables. For analyses of all endpoints, missing data for any reason was not imputed. If an outcome was missing, the subject was excluded from summary statistics and statistical analyses. If one of the biomarker measurements was missing, the subject was still considered for analysis if the particular predictive model did not include that biomarker as a variable.

To build a predictive model for each outcome, a subset of pre-selected potential factors was tested in a logistic model. These pre-identified factors were evaluated for inclusion using a randomly selected balanced dataset (50%) from GUARD-AKI. Expression of p14, p16, CD28, CD244, LAG3, suPAR (and their 2nd degree interaction), and sCr were evaluated. Using random sampling with replacement (>=200 iterations), logistic models were created using forward selection with model inclusion criteria of p-value<=0.25 after forced inclusion of p14 and p16. The percentages of inclusion for each of the factors was calculated. Factors with percentage approximately 4% were considered for further model retention and final model parameter estimates were derived using the entire sample.

The performance of each risk model was assessed by performing ROC analysis and measuring the total area under curve (AUC). A composite factor was created using each risk model and a distribution of values was generated across the complete sample. Probabilities of predicting each event were also modeled categorically in tertiles, with the lowest tertile serving as a reference group to derive an odds ratio for each outcome. The models were also adjusted for age, diabetes, CKD and CHF as potential effect modifiers.

Optimal thresholds for each model were identified by examining a distribution of fitted probabilities vs classification of the outcome in question. Sensitivity, specificity, PPV, NPV and accuracy were calculated at a threshold identified for each outcome model. Confidence intervals for sensitivity, specificity and accuracy are “exact” Clopper-Pearson confidence intervals and confidence intervals for PPV and NPV are standard logit confidence intervals as escribed by Mercaldo et al ^6^. Two-tailed p values of less than 0.5 were considered statistically significant. Statistical analyses were performed in SAS version 9.4 and JMP 12.2.0 (SAS Institute, Cary, NC).

## RESULTS

### Proof of concept of clinical relevance of biomarkers of senescence for risk prediction

To determine if measurements of senescence can identify patients at risk for adverse events after cardiac surgery, we measured expression of p16^INK4a^ (p16) mRNA in peripheral blood T cells, a master regulator and well-established biomarker of senescence^1, 7^, as well as the related transcript, p14^ARF^ (p14). Expression of p16 and p14 was measured pre-operatively in a cohort of 60 patients who underwent elective cardiac surgery at two centers. Study flow diagram and patient characteristics are shown in Figure S1 and Table S1. Given the small sample size, we focused on the most common adverse event post cardiac surgery, AKI. In this cohort, 30% (18/60) of patients developed in-hospital AKI.

**Table 1.** Baseline characteristics and postoperative course of GUARD-AKI study participants.

|  | Entire Cohort<br>N=331 | Site 1<br>N=160 | Site 2<br>N=136 | Site 3<br>N=35 |
| --- | --- | --- | --- | --- |
| **Demographics** |  |  |  |  |
| Age |  |  |  |  |
| mean (sd) | 67.2 (10) | 66.2 (9.3) | 69.1 (10.3) | 64.8 (10.7) |
| range | 43-91 | 43-91 | 43-87 | 45-84 |
| Male gender, n (%) | 262 (79.15) | 121 (75.63) | 114 (83.82) | 27 (77.14) |
| Race, n (%) |  |  |  |  |
| White | 261 (78.85) | 133 (83.13) | 105 (77.21) | 23 (65.71) |
| Black | 28 (8.46) | 22 (13.75) | 1 (0.74) | 5 (14.29) |
| Hispanic | 10 (3.02) | 1 (0.63) | 8 (5.88) | 1 (2.86) |
| Asian | 23 (6.95) | 2 (1.25) | 18 (13.24) | 3 (8.57) |
| BMI, mean (sd), kg/m<sup>2</sup> | 28.9 (5.7) | 29.5 (6.3) | 28.2 (4.9) | 28.9 (4.9) |
| eGFR, mean (sd), mL/min/1.73m<sup>2</sup> | 82.9 (21.5) | 80.8 (21.8) | 86.1 (20.2) | 80.4 (23.7) |
| LVEF, mean (sd), % | 53.0 (10.0) | 51.3 (9.7) | 55.0 (10.1) | 53.0 (9.9) |
| **Medical history** |  |  |  |  |
| Hypertension, n (%) | 268 (80.97) | 132 (82.5) | 104 (76.47) | 32 (91.43) |
| Diabetes, n (%) | 140 (42.3) | 75 (46.88) | 46 (33.82) | 19 (54.29) |
| Congestive heart failure, n (%) | 95 (28.7) | 67 (41.88) | 26 (19.12) | 2 (5.71) |
| Chronic kidney disease, n (%) | 58 (17.5) | 31 (19.4) | 18 (13.2) | 9 (25.7) |
| Atrial fibrillation, n (%) | 46 (13.9) | 20 (12.5) | 23 (16.9) | 3 (8.6) |
| Prior cardiac surgery, n (%) | 6 (1.81) | 1 (0.63) | 5 (3.68) | 0 (0) |
| ACE/ARB use, n (%) | 175 (53.03) | 89 (55.97) | 69 (50.74) | 17 (48.57) |
| **Procedural characteristics** |  |  |  |  |
| Urgent surgery, n (%) | 158 (47.73) | 78 (48.75) | 72 (52.94) | 8 (22.86) |
| Elective surgery, n (%) | 173 (52.27) | 82 (51.25) | 64 (47.06) | 27 (77.14) |
| Isolated CABG procedure, n (%) | 288 (87.01) | 143 (89.38) | 113 (83.09) | 32 (91.43) |
| 3+ vessels, n (%) | 257 (77.6) | 130 (81.3) | 99 (72.8) | 28 (80.0) |
| CPB time, mean (sd), min | 104.1 (42.2) | 107.6 (36.3) | 100.4 (47.5) | 102.7 (44.8) |
| ACE/ARB discontinuation, n (%) | 161 (92.53) | 89 (98.89) | 57 (82.61) | 15 (100) |
| **Post-operative course** |  |  |  |  |
| Time to extubation, h, mean (sd) | 9.3 (13.1) | 9.8 (3.9) | 8.3 (19.6) | 10.8 (8.5) |
| IABP, n (%) | 4 (1.21) | 3 (1.89) | 1 (0.74) | 0 (0) |
| ECMO, n (%) | 13 (3.93) | 2 (1.25) | 10 (7.35) | 1 (2.86) |
| Reoperation, n (%) | 9 (2.72) | 4 (2.5) | 2 (1.47) | 3 (8.57) |
| New onset Afib, n (%) | 101 (30.51) | 52 (32.5) | 37 (27.21) | 12 (34.29) |
| Transient Ischemic attack, n (%) | 1 (0) | 0 (0) | 1 (0) | 0 (0) |
| Cardiac arrest, n (%) | 3 (0) | 1 (0) | 1 (0) | 1 (0) |
| Acute Kidney Injury, n (%) |  |  |  |  |
| All stages | 66 (19.94) | 31 (19.38) | 30 (22.06) | 5 (14.29) |
| Stage 2 or above | 7 (2.1) | 4 (2.5) | 2 (1.47) | 1 (2.86) |
| Need for RRT, n (%) | 3 (0.91) | 2 (1.25) | 0 (0) | 1 (2.86) |
| New onset ESRD, n (%) | 2 (0.6) | 0 (0) | 2 (1.47) | 0 (0) |
| LOS ICU, mean (sd), h | 46.1 (46.9) | 52.8 (39.1) | 37.1 (54.7) | 50.0 (42.1) |
| LOS Hospital, mean (sd), d | 6.9 (6.2) | 7.4 (4.6) | 6.4 (8.0) | 7.0 (3.8) |
| Hospital death, n (%) | 3 (0.91) | 2 (1.25) | 0 (0) | 1 (2.86) |
| Discharge destination, n (%) |  |  |  |  |
| Home | 299 (91.16) | 148 (93.67) | 121 (88.97) | 30 (88.24) |
| Skilled nursing facility | 14 (4.27) | 4 (2.53) | 9 (6.62) | 1 (2.94) |
| Inpatient rehab | 15 (4.57) | 6 (3.8) | 6 (4.41) | 3 (8.82) |

A regression model that included p16, p14, the p16*p14 interaction, and pre-operative sCr could identify patients at risk for AKI with AUC of 0.76, 80% accuracy, and 86% NPV (Figure S2). Thus, we found that cellular senescence biomarkers measured prior to surgery have the capacity to predict the most common cardiac surgery-associated adverse event, AKI.

### Biomarker network used to characterize cellular senescence

While expression of p16 is a gold-standard measure of senescent cell load, and is a marker of established senescent cells ^7, 8^, accumulation of senescent cells depends on both formation of senescent cells and their clearance by the immune system ^9–11^ (Figure 1A). Broadly, with aging there is an increase in the rate of formation of senescent cells, and a decline in immune surveillance capacity, leading to a progressive accumulation of senescent cells^9, 10, 12^. Thus, measuring biomarkers of immune function in addition to p16 would allow us to better capture potential age-related vulnerability to adverse events, challenges associated with managing inflammatory responses induced by cardiac surgery, and overall capacity to recover.

**Figure 1.**
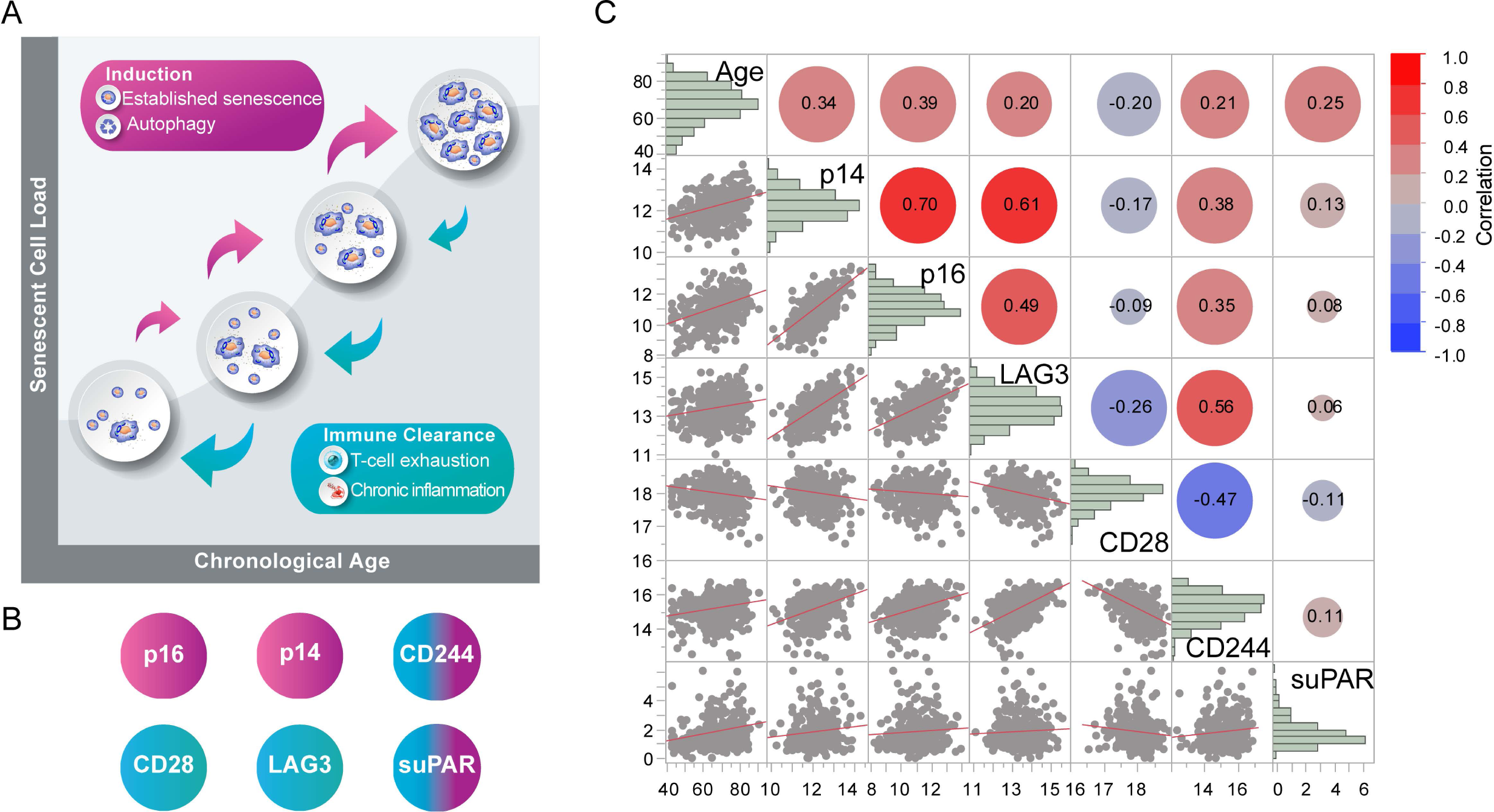
Cellular senescence network. A. Overall cellular senescence load is an outcome of two competing biological processes: induction of senescent cells due to cellular stress (pink arrows) and clearance of senescent cells by the immune system (teal arrows). B. Biomarkers of cellular senescence network used in this study. Biomarkers with primary known function in establishing senescence are shown in magenta, and those with immune system function are in teal. C. Scatterplot correlation matrix of pre-operative levels of cellular senescence network biomarkers as well as chronological age. The color of each correlation circle represents the correlation between each pair of variables on a scale from red (+1) to blue (−1). The size of each circle represents the significance test between the variables. A larger circle indicates a more significant relationship, and the Pearson correlation coefficient is shown as a number and a line of linear fit on the corresponding scatterplot. The histograms (diagonal across the matrix) show the distribution of each biomarker in the entire cohort.

The biomarkers shown in Figure 1 were selected for analysis based on studies in donors and patients in multiple clinical settings (manuscript in preparation), in order to capture established cellular senescence as well as age-dependent components of the adaptive and innate immune system. Briefly, CD28 and LAG3 are established markers of T cell exhaustion^13, 14^ . CD244 was first described as an exhaustion marker and has also been shown to correlate with age-dependent impairment of T cells^15^ ; however, more recently it was shown to regulate autophagy^16^, an important process that may also involve p14^17^. Finally, suPAR has been shown to be secreted by senescent cells^18, 19^; it is an immune-derived pathogenic factor of kidney disease and potentially cardiovascular disease^20, 21^. As expected, there is a large degree of association between these markers (Figure 1C) and a general, although weak, positive association with chronological age. CD28 expression declines with age consistent with a negative association with age and senescence markers. suPAR has the weakest association with senescent biomarkers consistent with the idea that total plasma levels reflect various sources of suPAR.

### GUARD-AKI study

A total of 331 participants who underwent cardiac surgery at three different centers were enrolled into the study and used in the analyses in this manuscript (Figure 2). Baseline characteristics of the patients in the entire cohort and at each site are shown in Table 1. The average age was 67 ± 10, 79% of the participants were male, and most participants (79%) were white. Mean BMI was 28.9, eGFR 82.9, LVEF 53; 81.0% had hypertension, 42.3% had diabetes, 28.7% had congestive heart failure, and 17.5% had chronic kidney disease. Surgery was elective for 52.3% patients and urgent for 47.7% (emergency surgery was an exclusion criterion). 87.0% of procedures were isolated CABG and 77.6% involved three or more vessels. Expression of biomarkers of senescence was measured pre-operatively and is shown in Table 2.

**Figure 2.**
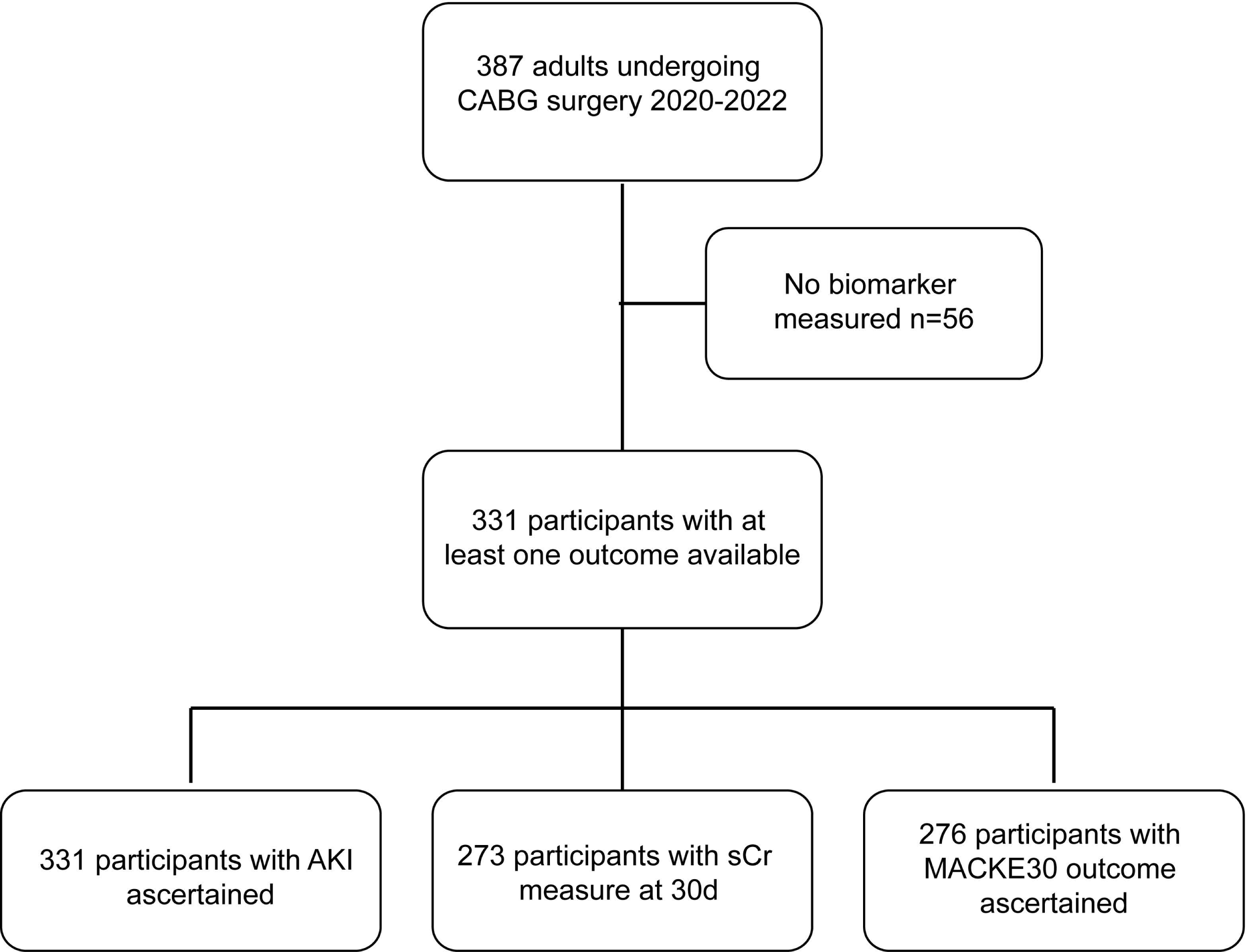
CONSORT flow diagram for GUARD-AKI study.

**Table 2.** Pre-operative levels of biomarkers of cellular senescence

|  | Entire Cohort | Site 1 | Site 2 | Site 3 |
| --- | --- | --- | --- | --- |
| p14, log2, mean (sd)* | 12.2 (0.7) | 12.3 (0.7) | 12.1 (0.7) | 12.2 (0.6) |
| p16, log2, mean (sd)* | 11.1 (1.1) | 11.4 (1.0) | 10.6 (1.1) | 10.9 (1.0) |
| LAG3, log2, mean (sd)* | 13.4 (0.8) | 13.7 (0.8) | 13.3 (0.8) | 13.2 (0.7) |
| CD28, log2, mean (sd)* | 18.0 (0.4) | 17.9 (0.4) | 18.0 (0.3) | 18.0 (0.4) |
| CD244, log2, mean (sd)* | 15.2 (0.8) | 15.4 (0.8) | 15.0 (0.6) | 15.1 (0.8) |
| suPAR ng/mL, mean (sd) | 1.8 (1.0) | 1.9 (1.2) | 1.5 (0.7) | 1.9 (0.8) |

\*arbitrary units, see Methods

### Preoperative biomarkers of cellular senescence predict AKI

AKI occurred in 20% of patients postoperatively. To determine if pre-operative biomarkers of cellular senescence can predict the incidence of AKI, we performed regression analyses that included biomarkers of cellular senescence, their pairwise interactions, and serum creatinine (Figure 3). The resulting model (see Methods for detailed description) had an AUC of 0.76. The cut-off for determining patients at risk for AKI was chosen to balance false positives and false negatives. At a cut-off of 30% probability, our model could identify patients at risk for AKI with 78.6% accuracy, 86.7% specificity, and 86.6% NPV.

**Figure 3.**
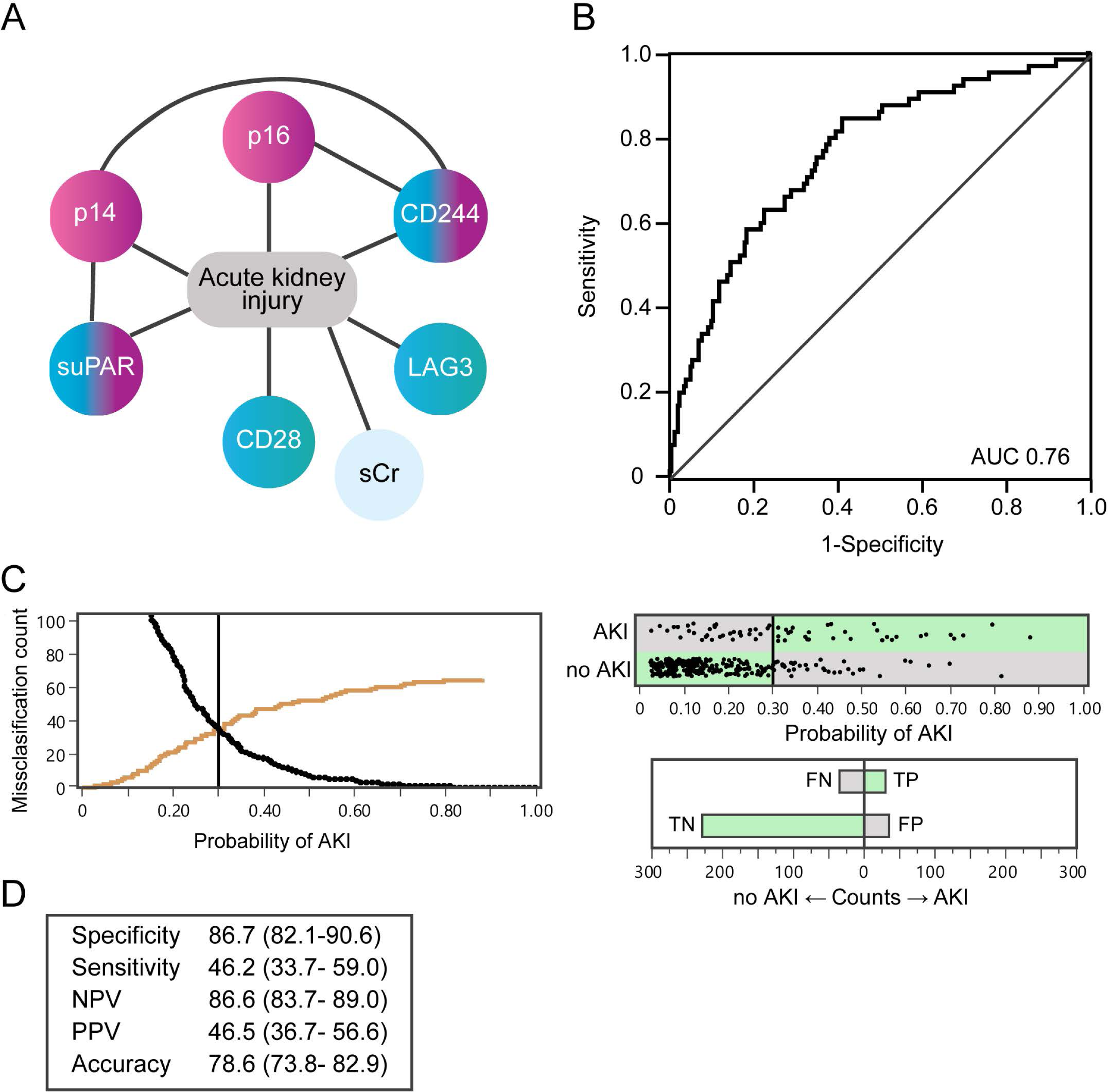
Cellular senescence-based model predicts cardiac surgery-associated AKI. A. Biomarkers of cellular senescence, their interactions (connecting lines), and sCr comprise the predictive AKI model. B. ROC analysis of the model. C. Threshold selection. Probability of AKI, derived from the predictive model, was plotted against AKI count to visualize patients at high risk for AKI (black line) vs low risk (brown line) (left panel). The vertical line shows the chosen probability threshold. Graphs to the right show the confusion matrix of actual vs. predicted AKI based on this threshold. Green bars are true and gray bars are false classifications. D. Performance of the AKI prediction model at the chosen threshold (value and 95% CI). NPV-negative predictive value, PPV-positive predictive value.

### Preoperative biomarkers of senescence predict decline in renal function at 30 days

Patient characteristics at a 30-day post-operative follow-up vist are shown in Table 3. Eleven percent of patients had a decline in kidney function compared to pre-operative status (decline in eGFR ≥25%). In addition, 13.4% had at least one major adverse cardiac and kidney event at the 30-day post-operative follow-up.

**Table 3.**
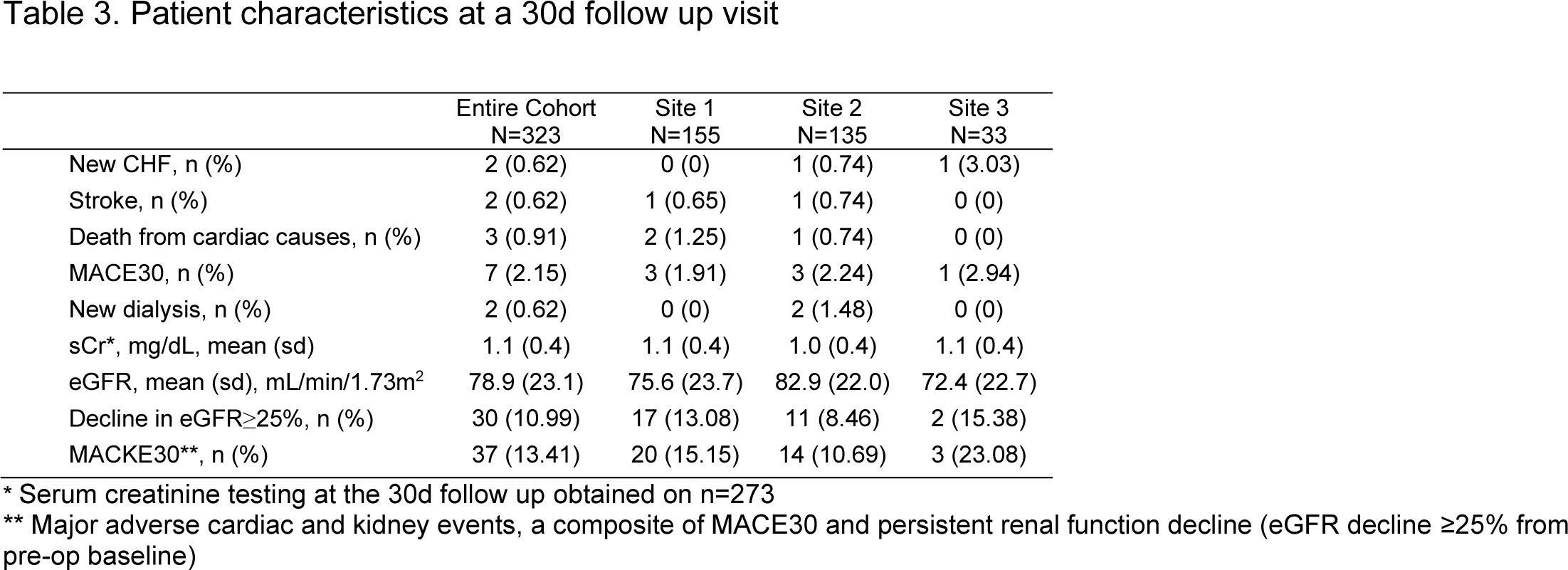
Patient characteristics at a 30d follow up visit

Post-discharge decline in eGFR is an important endpoint because there is substantial evidence that some patients who experience AKI will never recover their renal function and will experience an AKI-to-CKD progression. To determine if biomarkers of cellular senescence could predict the incidence of decline in eGFR at 30 days, we performed regression analyses as described for the AKI model, thereby generating another risk-prediction model with an AUC of 0.73 (Figure 4). For this model, the cut-off for designating patients at risk of renal decline was established to minimize false negatives. At a cut-off of 18% probability, our model could identify patients at risk for decline in renal function with 85.2% accuracy, 89.6% specificity, and 93.5% NPV.

**Figure 4.**
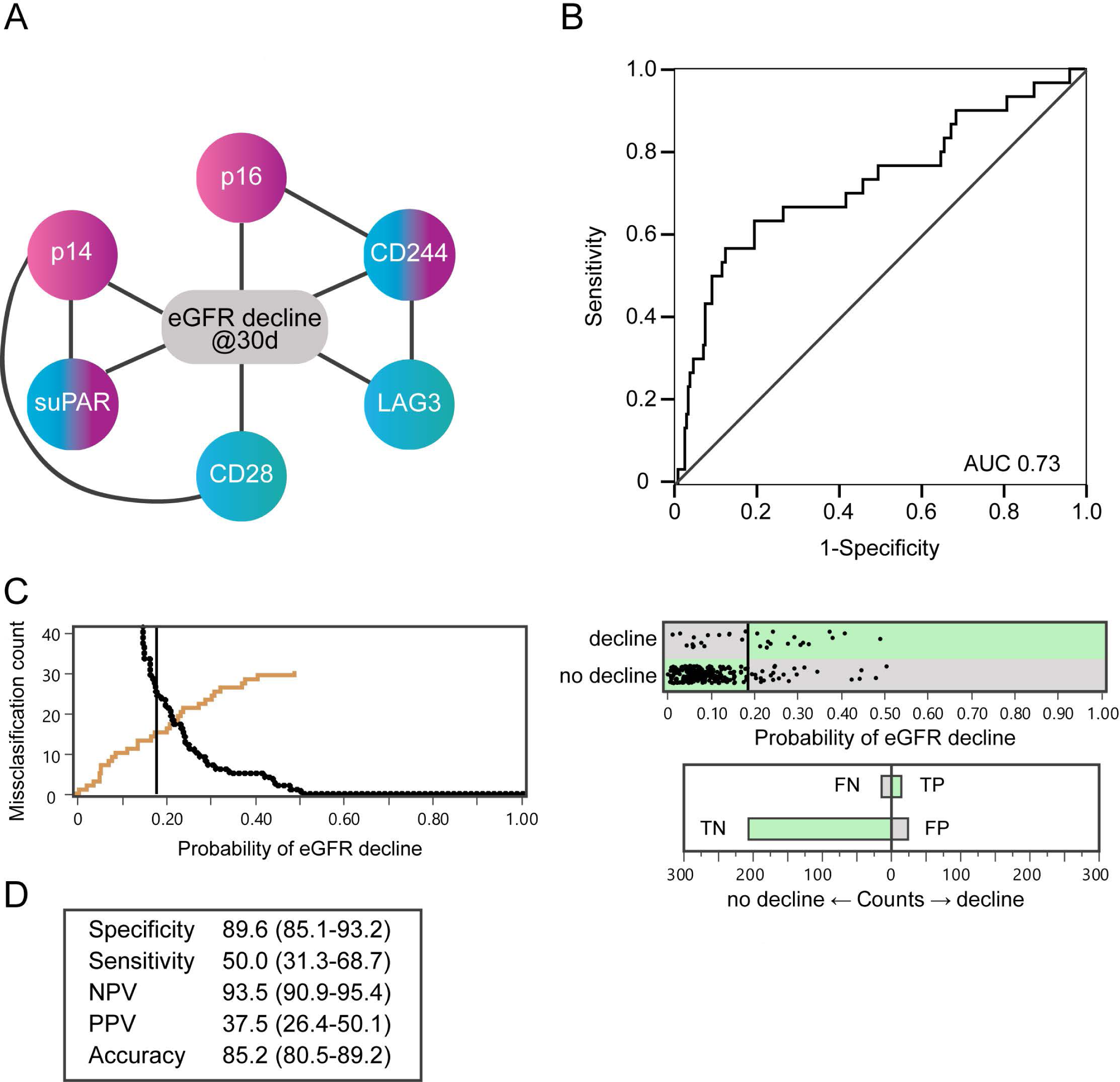
Cellular senescence-based model to predict decline in eGFR at a 30d post-operative follow-up visit. A. Biomarkers of cellular senescence and their interactions (connecting lines) comprise the predictive eGFR decline model. B. ROC analysis of the model. C. Threshold selection. Probability of eGFR decline, derived from the predictive model, was plotted against eGFR decline counts to visualize patients at high risk for eGFR decline (black line) vs. low risk (brown line) (left panel). The vertical line shows the chosen probability threshold. Graphs on the right show the confusion matrix of actual vs. predicted eGFR decline. Green bars are true and gray bars are false classifications. D. Performance of the eGFR decline prediction model at the chosen threshold (value and 95%CI).

### Incidence of AKI is largely non-overlapping with incidence of eGFR decline at 30 days

While these models demonstrated that biomarkers of senescence can predict patients at risk for both AKI and persisting eGFR decline, we noticed a difference in the components between the two models, including elimination of sCr as a risk factor in the eGFR decline model. When examined closely, we found that only 21% of patients who had in-hospital AKI also had a decline in eGFR at 30 days. Surprisingly, the majority (63%) of patients who had decline in eGFR did not have AKI during their hospital stay.

To better understand this phenomenon, we compared patient characteristics, multi-morbidities, and surgical factors across the AKI and eGFR decline endpoints. As expected, the incidence of AKI was strongly associated with known AKI risk factors such as age, CKD, hypertension, PVD, need for blood products post-operatively, and complex surgery (CABG+valve) as well as biomarkers of kidney inflammation (Activin A, sTNF-R1, and suPAR) (Table 4). Surprisingly, none of these risk factors for AKI was significantly associated with the incidence of eGFR decline. Moreover, while pre-operative CKD was strongly associated with AKI, it was not associated with post-operative eGFR decline and 80% of patients showing this eGFR decline did not have pre-existing CKD. This finding may explain why sCr was retained in the AKI prediction model but did not contribute to the eGFR decline prediction model. Thus, for most patients with eGFR decline at 30 days after surgery, this adverse event represents a new loss of kidney function, and not an AKI-to-CKD progression.

**Table 4.** Patient and surgical characteristics that are associated with risk of AKI are not associated with risk of eGFR decline at 30d. Only patients with ascertained outcomes for both AKI and 30d eGFR decline were included in the analysis.

|  | no AKI<br>n=220 | AKI<br>N=53 | p | no eGFR<br>decline<br>n=243 | eGFR<br>decline<br>n=30 | p |
| --- | --- | --- | --- | --- | --- | --- |
| **Demographic and clinical variables** |  |  |  |  |  |  |
| Age, mean (sd) | 67 (10) | 71 (9) | **0.004\*** | 67 (10) | 70 (9) | 0.13 |
| Male, n (%) | 174 (79) | 40 (75) | 0.57 | 193 (79) | 21 (70) | 0.24 |
| Pre-op eGFR, mean (sd) | 85 (20) | 72 (24) | **0.0001\*** | 83 (21) | 83 (20) | 0.9 |
| CKD, n (%) | 28 (13) | 20 (38) | **0.0001\*** | 42 (17) | 6 (20) | 0.71 |
| Hypertension, n (%) | 171 (78) | 47 (89) | 0.07** | 194 (80) | 24 (80) | 0.98 |
| CHF, n (%) | 64 (29) | 16 (30) | 0.87 | 69 (28) | 11 (37) | 0.35 |
| Atrial fibrillation | 33 (12) | 13 (20) | 0.13 | 36 (15) | 4 (13) | NA<sup>1</sup> |
| MI, n (%) | 71 (32) | 23 (43) | 0.13 | 83 (34) | 11 (37) | 0.78 |
| PVD, n (%) | 8 (94) | 7 (83) | **0.02\*** | 13 (5) | 2 (7) | NA<sup>1</sup> |
| Diabetes, n (%) | 89 (40) | 25 (47) | 0.37 | 106 (44) | 8 (27) | 0.08** |
| Insulin-depend diabetes, n (%) | 33 (15) | 12 (23) | 0.18 | 40 (16) | 5 (17) | 0.98 |
| CABG +valve surgery, n (%) | 25 (11) | 11 (21) | 0.07** | 31 (13) | 5 (17) | 0.55 |
| Use of post-op blood products | 39 (18) | 19 (36) | **0.004\*** | 51 (21) | 7 (23) | 0.77 |
| Post-op NOAF, n (%) | 59 (27) | 21 (40) | 0.07** | 60 (25) | 20 (67) | **0.0001\*** |
| **Pre-op kidney biomarkers** |  |  |  |  |  |  |
| Activin A, pg/ml, mean (sd)<sup>2</sup> | 185 (138) | 252.9 (210.6) | **0.02\*** | 205 (174) | 182 (80) | 0.69 |
| sTNF-R1, pg/ml, mean (sd)<sup>3</sup> | 1360 (775) | 1849(1019) | **0.004\*** | 1462 (832) | 1547 (642) | 0.14 |
| suPAR, ng/ml, mean (sd)<sup>4</sup> | 1.7 (0.9) | 2.4 (1.3) | **0.0001\*** | 1.8 (1.0) | 2.0 (1.1) | 0.51 |

\* p<0.05, \*\* p<0.1
<sup>1</sup> less than 5 events per group
<sup>2</sup> 193 samples that had AKI endpoint and 155 samples that had a 30d eGFR decline endpoint were used for Activin A testing
<sup>3</sup> 165 samples that had AKI endpoint and 128 samples that had a 30d eGFR decline endpoint were used for TNFR1 testing
<sup>4</sup> 329 samples that had AKI endpoint and 271 samples that had a 30d eGFR decline endpoint were used for suPAR testing

Interestingly, incidence of new onset post-surgical atrial fibrillation (NOAF) was highly associated with the incidence of eGFR decline at 30 days, but only weakly associated with incidence of AKI. And while not statistically significant, patients with congestive heart failure were enriched in the group with eGFR decline as compared to the group with no eGFR decline (37% vs. 28%), but equally distributed in patients with or without AKI (30% vs. 29%). Together, these results suggest different etiologies for these adverse events.

### Preoperative biomarkers of senescence predict a composite of cardiac and kidney events at 30 days

Given the interdependent nature of heart and kidney function and disease, a composite of cardiac and kidney adverse events at 30 days post-surgery (MACKE30) was tested as an outcome. 13.4% patients had a MACKE30 event. Using the same markers and methods from the previous two models, we built a third regression analysis model with an AUC of 0.71. Similar to the model for eGFR decline prediction, the cut point was established to minimize false negatives (Fig. 5). At the cut-off of 19% probability, our model could identify patients at risk for MACKE30 with 79.9% accuracy, 84.8% specificity, and 91.4% NPV.

**Figure 5.**
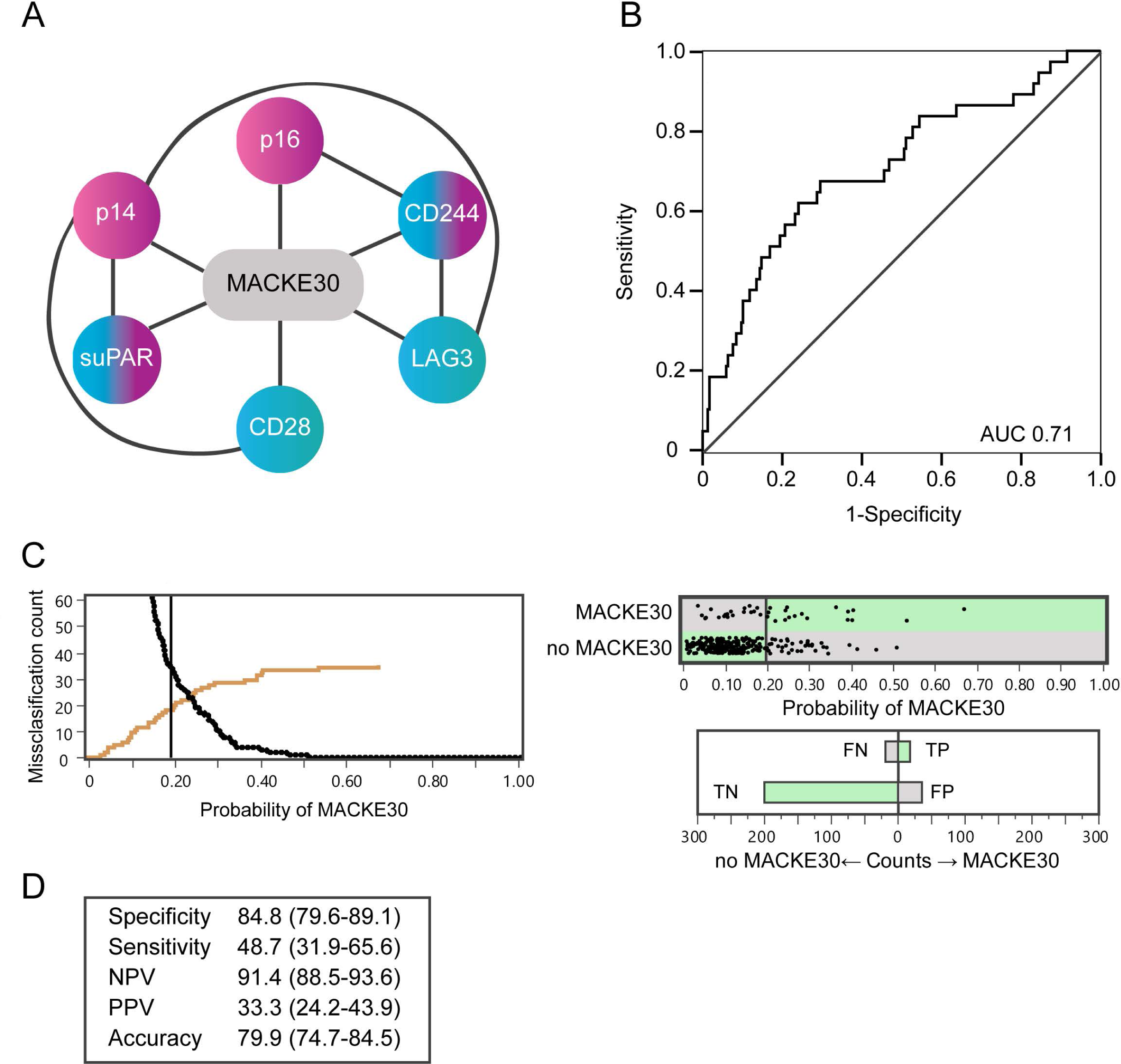
Cellular senescence-based model to predict major adverse cardiac and kidney events at the 30d post-op (MACKE30). A. Biomarkers of cellular senescence and their interactions (connecting lines) that comprise the predictive MACKE30 model. B. ROC analysis of the model. C. Threshold selection. Probability of MACKE30, derived from the predictive model, was plotted against MACKE30 counts to visualize patients at high risk for MACKE30 (black line) vs. low risk (brown line) (left panel). The vertical line shows the chosen probability threshold. Graphs on the right demonstrate the confusion matrix of actual vs. predicted MACKE30. Green bars are true and gray bars are false classifications. D. Performance of the MACKE30 prediction model at the chosen threshold (value and 95%CI).

### Models based on senescence biomarkers are not improved by patient characteristics

To assess the value of using cellular senescence biomarkers relative to demographics and multi-morbidities that are commonly used in patient care, we determined both the odds ratio of having an event based on a biomarker model, and the adjusted odds ratio when clinical variables were added to the model. Predicted probability of an event was examined in tertiles and the probabilities in the highest tertile was compared to those in the lowest tertile. Patients in the highest tertile of the cellular senescence biomarker-based model of AKI had 7.8 (95%CI 3.3-18.4, p=0.0001) higher odds of developing AKI than patients in the lowest tertile (Figure 6). Adjustment for clinical variables decreased the average odds to 5.5 (95%CI 2.2-13.7), but AKI prediction remained highly statistically significant (p=0.0003). A similar approach was used to estimate odds of decline in eGFR and MACKE30 using non-adjusted and adjusted senescence biomarker-based models. Again, senescence-biomarker-based models remained highly statistically significant but largely unchanged after the adjustment. These data suggest that biomarkers of cellular senescence are identifying patients at risk for cardiac surgery-associated adverse events independently of clinical factors.

**Figure 6.**
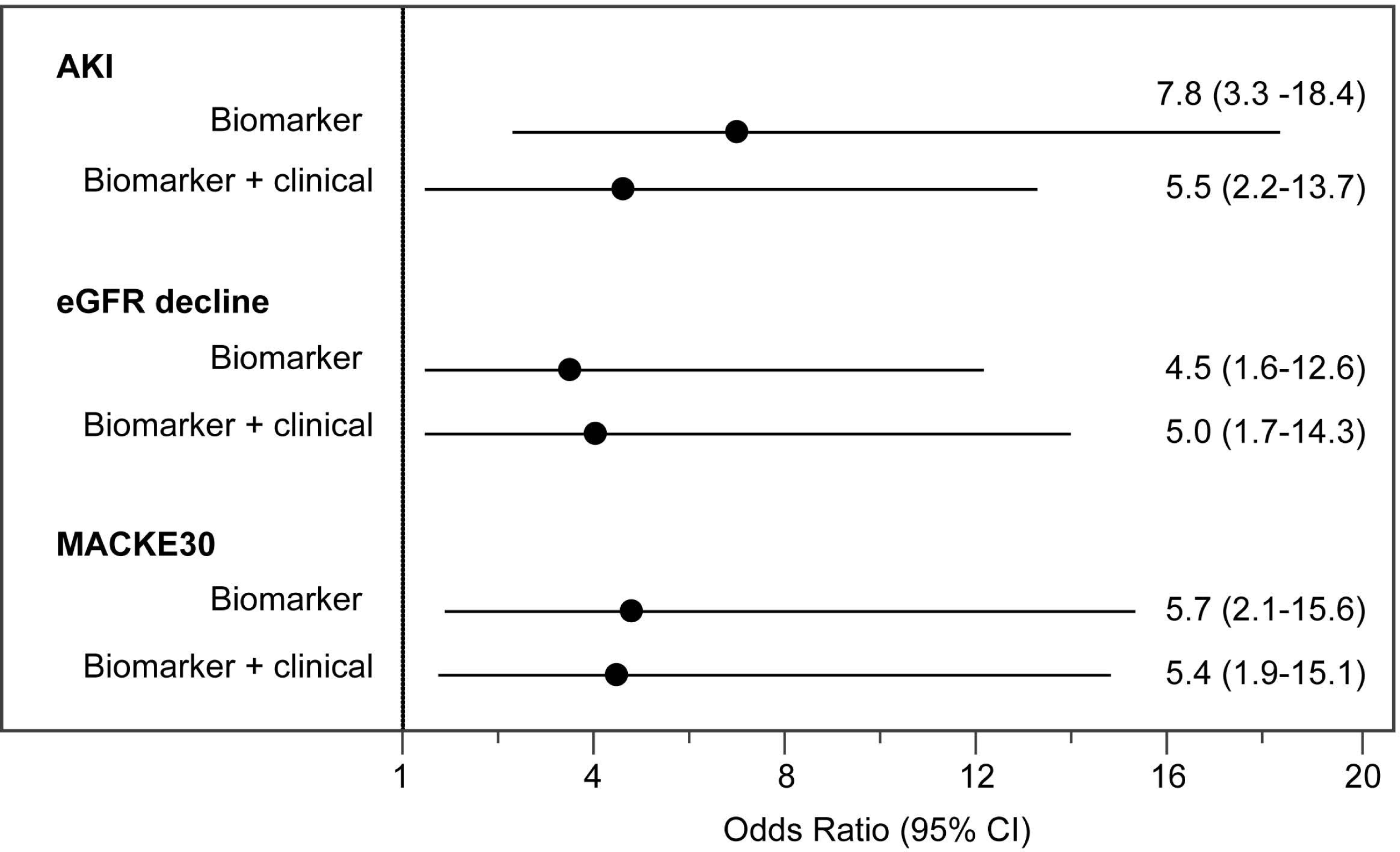
Predictive ability of senescence-based models is unchanged after adjustment for clinical variables. Odds ratios, with 95% CI, are plotted for cellular senescence-based models (Biomarker) for AKI, 30d decline in eGFR or MACKE30. Each model was also adjusted for age, CKD, diabetes and CHF (Biomarker + clinical).

## DISCUSSION

This is the first report using pre-operative, aging-related biomarkers of cellular senescence and immune system function to predict risk of common and serious cardiac surgery-related adverse events including in-hospital AKI, a decline in eGFR by ≥25% at 30 days, and a composite of cardiac and kidney adverse events at 30 days.

Biomarkers of aging represent a significant conceptual departure from other measures of patient risk that, historically, have been disease or organ-specific, and thereby fail to capture the multi-system vulnerability associated with aging. As a primary mechanism of aging, senescent cells increase in abundance over time. However, senescent cell load varies dramatically between individuals, including among those of the same chronological age^1, 7^. The rate of accumulation of senescent cells is a balance between senescence induction and clearance which is mediated by the immune system^9, 10, 12^. Induction of cellular senescence in immune cells alone was recently shown to induce senescence in tissues and organs throughout the body^22^, and there is increasing evidence that immune system impairment inhibits clearance of senescent cells and contributes to age-related conditions^11, 23^. The data presented here support the assertion that senescent cell load is a key, multi-system risk factor for adverse events after cardiac surgery. Senescence-based risk alone appears sufficient to stratify patients by risk of AKI, eGFR decline at 30 days and MACKE30. Patients in the highest tertile had between 4.5-7.8 higher odds of developing an outcome independently of chronological age, CKD, diabetes, and CHF.

Given the interdependent and perpetuating nature of kidney and cardiac function, there has been considerable effort in the past to predict risk of AKI. The only available molecular diagnostic, NephroCheck, uses kidney-specific biomarkers to identify injury after it occurs, eliminating the opportunity for prevention and the earliest intervention^24^. Predictive algorithms based on clinical variables alone have focused on only the most severe outcomes, such as the Thakar score^25^, or they have utilized intra-operative and post-operative variables that preclude use in everyday practice. In contrast, the biomarkers used in this study are measured pre-operatively and can be integrated into clinical decision making throughout the peri-operative period. Further, cellular senescence biomarkers are not kidney-specific, but rather capture overall organismal aging as well as immune system status. The multi-system nature of aging vulnerability captured here underlies the success of this study. Measuring the senescence network in the immune system has the potential to define the physiologic impact of senescent cell load on age-related dysfunction, low grade chronic inflammation, and reduced capacity to heal.

We also observed that there is only 21% overlap between the patients who experienced AKI and those with a ≥25% decline in eGFR at 30 days, suggesting different etiologies. New post-operative eGFR decline, in the absence of prior renal decline (AKI or CKD), may instead associate more closely with cardiac dysfunction, since the population is enriched with those experiencing new onset atrial fibrillation, and potentially a history of congestive heart failure. Given that the vast majority of patients (80%) experiencing eGFR decline at 30 days did not have CKD (pre-op eGFR <60 ml/min/1.73m^2^), they may not currently be seen as high risk for kidney injury.

Early, pre-operative identification of risk through senescence biomarker testing both reveals unexpected risk and allows for careful targeting of peri-operative interventions, such as the recently proposed order set for preventing AKI^26^. These interventions include intensive fluid management and hemodynamic monitoring, limited nephrotoxin exposure, and strict metabolic management. In addition, patients with low-risk biomarker status could be identified as candidates for an accelerated recovery plan, with potential for earlier mobility, line removal, transfer to less intensive care, NSAID vs opioid pain management, and early discharge. The predictive models in this study were designed with high specificity and negative predictive values to prioritize true negatives, i.e., higher confidence in “fast track” recovery status, at the cost of more false positives, i.e., utilization of a prevention focused order set in some patients who are not at-risk. This approach is well-aligned with the rapidly expanding ERAS Cardiac movement which has demonstrated that multi-modal, peri-operative interventions can improve patient outcomes, meet quality goals, and improve patient and staff satisfaction^3, 27^.

Beyond the discovery nature of this study, other limitations include the limited number of sites and enrollment during the pandemic. The characteristics of patients undergoing surgery may have been affected, whether through changing hospital policies or by changing preferences among both providers and patients regarding selection and timing of surgery. Future studies will be conducted to validate these predictive models, demonstrate clinical utility, and develop a molecular prognostic that can be utilized in everyday surgical practice.

In summary, this is the first report utilizing senescence biomarkers for risk prediction to pre-operatively identify cardiac surgery patients at-risk of adverse events. These findings lay the foundation for future studies that can bring molecular aging to pre-surgical risk assessment, improving both clinical outcomes and resource utilization in cardiac surgery.

## Data Availability

Due to the informed consent and data privacy policies, the clinical data are not publicly available, i.e., accessible for anyone, for any purpose without a review by the Central IRB on a project-by-project basis. Requests for raw data can be made to the corresponding author.

## LIST OF TABLES

## LIST OF FIGURES

## ACKNOWLEDGMENTS

We would like to acknowledge surgeons and clinical research staff who worked tirelessly to make this study possible: Trevor Upham, MD, Gene Graham, Taylor Terry, Rhonda Norton, Taylor Guidi, Michelle Parish, Tiffany Bisanar, and Michael Smith.

This work was funded in part by grants from the NIH/NIA (R43AG050353 and R44AG060888). The funding sources had no involvement in the study design or in the collection, analysis and interpretation of data. The funding sources also had no involvement in the writing of this report or in the decision to submit the report for publication.

## REFERENCES

1. He S, Sharpless NE. Senescence in Health and Disease. Cell. 2017;169(6):1000–1011.

2. Calcinotto A, Kohli J, Zagato E, Pellegrini L, Demaria M, Alimonti A. Cellular senescence: Aging, cancer, and injury. Physiological Reviews. 2019;99(2):1047–1078.

3. Williams JB, Mcconnell G, Allender JE, et al. One-year results from the first US-based enhanced recovery after cardiac surgery (ERAS Cardiac) program. The Journal of Thoracic and Cardiovascular Surgery. 2019;157:1881–1888.

4. Levey AS, Stevens LA, Schmid CH, et al. A new equation to estimate glomerular filtration rate. Ann Intern Med. 2009;150(9):604–612.

5. Mitin N, Nyrop KA, Strum SL, et al. A biomarker of aging, p16, predicts peripheral neuropathy in women receiving adjuvant taxanes for breast cancer. npj Breast Cancer. 2022;8(1):103.

6. Mercaldo ND, Lau KF, Zhou XH. Confidence intervals for predictive values with an emphasis to case-control studies. Stat Med. 2007;26(10):2170–2183.

7. Liu Y, Sanoff HK, Cho H, et al. Expression of p16 INK4a in peripheral blood T-cells is a biomarker of human aging. Aging Cell. 2009;8(4):439–448.

8. Kim WY, Sharpless NE. The regulation of INK4/ARF in cancer and aging. Cell. 2006;127(2):265–275.

9. Song P, An J, Zou MH. Immune Clearance of Senescent Cells to Combat Ageing and Chronic Diseases. Cells. 2020;9(3):671.

10. Pereira BI, Devine OP, Vukmanovic-Stejic M, et al. Senescent cells evade immune clearance via HLA-E-mediated NK and CD8+ T cell inhibition. Nature Communications. 2019;10(1).

11. Muñoz DP, Yannone SM, Daemen A, et al. Targetable mechanisms driving immunoevasion of persistent senescent cells link chemotherapy-resistant cancer to aging. JCI Insight. 2019;4(14).

12. Marin I, Serrano M, Pietrocola F. Recent insights into the crosstalk between senescent cells and CD8 T lymphocytes. Aging. 2023;9.

13. Goronzy JJ, Weyand CM. Understanding immunosenescence to improve responses to vaccines. Nat Immunol. 2013;14(5):428–436.

14. Rodriguez IJ, Lalinde Ruiz N, Llano León M, et al. Immunosenescence Study of T Cells: A Systematic Review. Frontiers in Immunology. 2021;11. Accessed April 3, 2023.

15. Wang X, Wang D, Du J, et al. High Levels of CD244 Rather Than CD160 Associate With CD8+ T-Cell Aging. Frontiers in Immunology. 2022;13. Accessed October 23, 2022.

16. Chaudhary A, Leite M, Kulasekara BR, et al. Human Diversity in a Cell Surface Receptor that Inhibits Autophagy. Current Biology. 2016;26(14):1791–1801.

17. Ozenne P, Eymin B, Brambilla E, Gazzeri S. The ARF tumor suppressor: Structure, functions and status in cancer. International Journal of Cancer. 2010;127(10):2239–2247.

18. Coppé JP, Patil CK, Rodier F, et al. Senescence-associated secretory phenotypes reveal cell-nonautonomous functions of oncogenic RAS and the p53 tumor suppressor. PLoS biology. 2008;6(12).

19. Franceschi C, Campisi J. Chronic inflammation (inflammaging) and its potential contribution to age-associated diseases. J Gerontol A Biol Sci Med Sci. 2014;69 Suppl 1:S4–9.

20. Hayek SS, Leaf DE, Samman Tahhan A, et al. Soluble Urokinase Receptor and Acute Kidney Injury. New England Journal of Medicine. 2020;382(5):416–426.

21. Hindy G, Tyrrell DJ, Vasbinder A, et al. Increased soluble urokinase plasminogen activator levels modulate monocyte function to promote atherosclerosis. J Clin Invest. 2022;132(24).

22. Yousefzadeh MJ, Flores RR, Zhu Y, et al. An aged immune system drives senescence and ageing of solid organs. Nature. 2021;594(7861):100–105.

23. Ovadya Y, Krizhanovsky V. Strategies targeting cellular senescence. Journal of Clinical Investigation. 2018;128(4):1247–1254.

24. Meersch M, Schmidt C, Van Aken H, et al. Urinary TIMP-2 and IGFBP7 as early biomarkers of acute kidney injury and renal recovery following cardiac surgery. PLoS ONE. 2014;9(3):1–9.

25. Thakar CV, Arrigain S, Worley S, Yared JP, Paganini EP. A clinical score to predict acute renal failure after cardiac surgery. J Am Soc Nephrol. 2005;16(1):162–168.

26. Engelman DT, Shaw AD. A Turnkey Order Set for Prevention of Cardiac Surgery–Associated Acute Kidney Injury. The Annals of Thoracic Surgery. 2023;115(1):11–15.

27. Engelman DT, Crisafi C, Germain M, Greco B, Nathanson BH, Engelman RM, Schwann TA. Using urinary biomarkers to reduce acute kidney injury following cardiac surgery. J Thorac Cardiovasc Surg. 2020 Nov;160(5):1235–1246.

