## Supplemental Information for "A signature of pre-operative biomarkers of cellular senescence to predict risk of cardiac and kidney adverse events after cardiac surgery"

**SUPPLEMENTAL DATA**

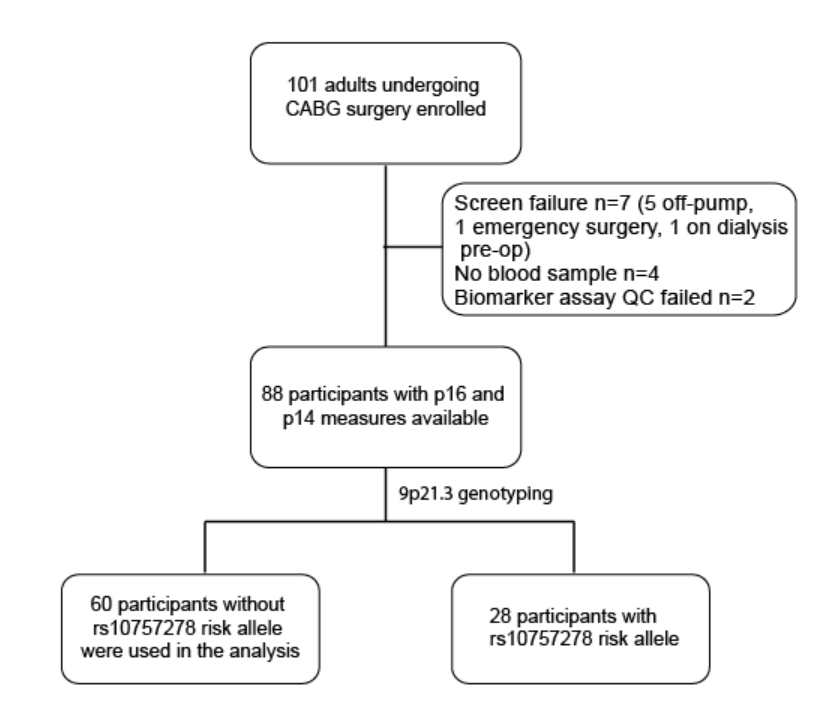

Figure S1. CONSORT flow diagram of pilot study. 88 participants that had biomarker data and the AKI endpoint available were tested for homozygous mutation at the rs10757278 risk allele on 9p21.3, a locus that encodes both p16 and p14. Participants who did not have a mutation at this risk allele were used in the analysis.

Table S1. Patient characteristics in the pilot study.

| **Variables** | **Pilot study n=60** |
| --- | --- |
| Age, mean (sd) | 64.8 (9.8) |
| Male gender, n(%) | 44 (75) |
| White, n(%) | 54 (90) |
| sCr, mean (sd) | 1.0 (0.3) |
| eGFR, mean (sd) | 75.8 (20.0) |
| log2 p16, a.u. mean (sd) | 9.5 (1.3) |
| log2 p14, a.u. mean (sd) | 12.5 (0.6) |
| EF, mean (sd) | 53 (9) |
| CKD, n(%) | 9 (15) |
| Diabetes, n(%) | 14 (23) |
| Isolated CABG, n(%) | 54 (90) |
| Post-op AKI, n(%) | 18 (30) |

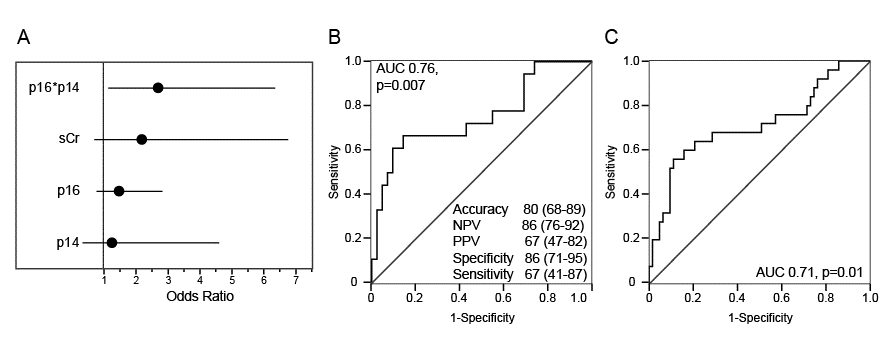

Figure S2. Prediction of cardiac surgery-associated AKI by biomarkers of cellular senescence. A. Composition of the model containing biomarkers of senescence and serum creatinine to predict AKI. B. ROC analysis and model performance. C. ROC analysis of the model to predict AKI containing the same variables but expanded to a population of 88 participants (not stratified by the mutational status at rs10757278).
